# Mapping physical access to healthcare for older adults in sub-Saharan Africa: A cross-sectional analysis with implications for the COVID-19 response

**DOI:** 10.1101/2020.07.17.20152389

**Authors:** Pascal Geldsetzer, Marcel Reinmuth, Paul O. Ouma, Sven Lautenbach, Emelda A. Okiro, Till Bärnighausen, Alexander Zipf

**Author notes:** Joint first authors (Pascal Geldsetzer, Marcel Reinmuth). **Corresponding author:** Pascal Geldsetzer MBChB ScD MPH Division of Primary Care and Population Health, Department of Medicine, Stanford University, Stanford, CA, USA. Full professors: Till Bärnighausen, Alexander Zipf.

## Abstract

**Background:** SARS-CoV-2, the virus causing coronavirus disease 2019 (COVID-19), is rapidly spreading across sub-Saharan Africa (SSA). Hospital-based care for COVID-19 is particularly often needed among older adults. However, a key barrier to accessing hospital care in SSA is travel time to the healthcare facility. To inform the geographic targeting of additional healthcare resources, this study aimed to determine the estimated travel time at a 1km x 1km resolution to the nearest hospital and to the nearest healthcare facility of any type for adults aged 60 years and older in SSA.

**Methods:** We assembled a unique dataset on healthcare facilities’ geolocation, separately for hospitals and any type of healthcare facility (including primary care facilities) and including both private- and public-sector facilities, using data from the OpenStreetMap project and the KEMRI Wellcome Trust Programme. Population data at a 1km x 1km resolution was obtained from WorldPop. We estimated travel time to the nearest healthcare facility for each 1km x 1km grid using a cost-distance algorithm.

**Findings:** 9.6% (95% CI: 5.2% – 16.9%) of adults aged ≥60 years had an estimated travel time to the nearest hospital of longer than six hours, varying from 0.0% (95% CI: 0.0% – 3.7%) in Burundi and The Gambia, to 40.9% (95% CI: 31.8% – 50.7%) in Sudan. 11.2% (95% CI: 6.4% – 18.9%) of adults aged ≥60 years had an estimated travel time to the nearest healthcare facility of any type (whether primary or secondary/tertiary care) of longer than three hours, with a range of 0.1% (95% CI: 0.0% – 3.8%) in Burundi to 55.5% (95% CI: 52.8% – 64.9%) in Sudan. Most countries in SSA contained populated areas in which adults aged 60 years and older had a travel time to the nearest hospital of more than 12 hours and to the nearest healthcare facility of any type of more than six hours. The median travel time to the nearest hospital for the fifth of adults aged ≥60 years with the longest travel times was 348 minutes (equal to 5.8 hours; IQR: 240 – 576 minutes) for the entire SSA population, ranging from 41 minutes (IQR: 34 – 54 minutes) in Burundi to 1,655 minutes (equal to 27.6 hours; IQR: 1065 – 2440 minutes) in Gabon.

**Interpretation:** Our high-resolution maps of estimated travel times to both hospitals and healthcare facilities of any type can be used by policymakers and non-governmental organizations to help target additional healthcare resources, such as new make-shift hospitals or transport programs to existing healthcare facilities, to older adults with the least physical access to care. In addition, this analysis shows precisely where population groups are located that are particularly likely to under-report COVID-19 symptoms because of low physical access to healthcare facilities. Beyond the COVID-19 response, this study can inform countries’ efforts to improve care for conditions that are common among older adults, such as chronic non-communicable diseases.

**Funding:** Bill & Melinda Gates Foundation

**Research in context:** 

**Evidence before this study:** We searched MEDLINE from January 1966 until May 2020 for studies with variations of the words ‘physical access’, ‘distance’, ‘travel time’, ‘hospital’, and ‘healthcare facility’ in the title or abstract. To date, the only studies to systematically map physical access to healthcare facilities in sub-Saharan Africa at a high resolution examined access to emergency hospital care (with a focus on women of child-bearing age), access to care for children with fever, travel time to the nearest healthcare facility for specific populations at risk of viral haemorrhagic fevers, and travel time to the nearest regional- or district-level hospital.

**Added value of this study:** The added value of this study is threefold. First, we assembled a new dataset of GPS-tagged healthcare facilities, which combines two unique data sources for the geolocation of healthcare facilities across sub-Saharan Africa: one-based on crowd-sourced data from OpenStreetMap and one based on information from ministries of health, health management information systems, government statistical agencies, and international organizations. Second, this is the first study to comprehensively map both hospitals and primary healthcare facilities, and including both public- and private-sector facilities, across sub-Saharan Africa. Third, because the COVID-19 epidemic causes a far higher need for hospital services among older than younger population groups, we focus on physical access to healthcare for the population aged 60 years and older, which is a population group that is rarely studied in investigations of healthcare demand and supply in the region. As such, our maps can inform not only the health system response to COVID-19, but more generally to conditions that are common among older adults in the region, particularly chronic non-communicable diseases and their sequelae.

**Implications of all the available evidence:** Low physical access to healthcare in sub-Saharan Africa will be a major barrier to receiving care for adults aged 60 years and older with COVID-19. However, there is a wide degree of variation in physical access to healthcare facilities for older adults in the region both between and within countries, which likely has an important bearing on the extent to which different population groups within countries are able to access care for COVID-19. Likewise, in those areas with a long travel time to the nearest healthcare facility of any type (which exist in most countries), symptomatic cases of COVID-19 are particularly unlikely to be reported to the healthcare system. Our high-resolution maps for each region and country in sub-Saharan Africa provide precise information about this geographic variation for local, national, and regional policymakers as well as non-governmental organizations.

## Introduction

Declared a pandemic by the World Health Organization on March 11 2020,^1^ SARS-CoV-2 has caused over nine million confirmed infections and the disease it can trigger – coronavirus disease 2019 (COVID-19) – has led to almost 500,000 reported deaths across the world by late June 2020.^2^ While low testing numbers do not allow for a reliable assessment of the extent of the epidemic in sub-Saharan Africa (SSA), the region had over 300,000 reported infections and almost 9,000 deaths due to SARS-CoV-2 as of June 24 2020.^2^ Epidemiological modelling suggests that COVID-19 could lead to between 300,000 and 2.5 million deaths in SSA, depending on modelling assumptions and the mitigation policies that are adopted.^3^

There are numerous barriers to receiving high-quality healthcare in SSA, including financial barriers to accessing care, weak supply chains, and understaffing of healthcare facilities.^4^ However, physical distance to the nearest healthcare facility – and the associated requirements for transport options, cost of transport, and time lost from other income-generating activities – consistently figure as one of the most important barriers to accessing both hospital-based and primary care in the region.^5-10^

Travel time to the nearest healthcare facility and the nearest hospital will likely also play an important role in the ability of health systems in SSA to respond to SARS-CoV-2 for three main reasons. First, physical access to hospitals will influence whether and how timely individuals with COVID-19 are able to seek healthcare. While many hospitals in SSA are not able to provide mechanical ventilation,^11,12^ other critical components of care for those with severe COVID-19, such as hemodynamic support, supplemental oxygen therapy, and treatment of coinfections (e.g., bacterial pneumonia), are more readily available in hospitals in SSA.^13,14,15^ Second, physical access to a healthcare facility of any type will impact whether and when during the disease course individuals with COVID-19 contact the healthcare system. These care-seeking decisions in turn have important ramifications for whether the health system is notified of COVID-19 cases and, thus, the monitoring of the epidemic, particularly in settings that are unable to conduct large-scale community-based testing for SARS-CoV-2 infections. Third, if these options become widely available in SSA in the future, physical access to healthcare facilities will likely affect to what degree individuals with COVID-19 take up effective medications against the condition and possibly also to what degree they are able to access a vaccine against SARS-CoV-2.

Having a detailed understanding of where population groups are located that are both vulnerable to COVID-19 and have long travel times to the nearest healthcare facility can inform where additional healthcare resources (e.g., the establishment of makeshift hospitals or programs to ensure availability of transport to hospitals) are most needed. In addition, such knowledge would allow for pinpointing those geographic areas that are most likely to harbour the most cases of COVID-19 that were unreported due to lack of physical access to care, which in turn can inform the geographic targeting of testing efforts. More broadly, understanding where older adults reside who have the least physical access to healthcare can inform health systems’ efforts to improve care for conditions that are common in this age group, particularly chronic non-communicable diseases and their sequelae. By assembling a unique dataset from both crowd-sourced data and official records by governments and international organizations, this study, therefore, aimed to create highly detailed maps of estimated travel time for adults aged 60 years and older in SSA to both the nearest hospital and the closest healthcare facility of any type.

## Methods

### Data sources for the geolocation of healthcare facilities

We used two data sources: healthcare facility data from the OpenStreetMap (OSM) project and a geocoded inventory of healthcare facilities published by the KEMRI Wellcome Trust Research Programme.^16,17^ OSM is a collaborative online platform to map, edit, and share geospatial data globally. Started in 2004, OSM evolved from a crowd-sourced alternative for proprietary map data providers, to an important complementary data source in humanitarian settings,^18,19^ and a widely used source of information for base maps as well as for critical infrastructure in the global South.^20^ Querying the database for all objects with either "amenity” or “healthcare” as key and either “hospital”, “clinic”, or “doctors” as value, we extracted all healthcare facilities mapped in OSM with their geographic coordinates using the ohsome api.^21^ We identified 24,571 healthcare facilities of which 13,392 were tagged as hospitals.

The second data source used in this analysis was an inventory of 98,745 public-sector healthcare facilities across all countries of SSA except for five small island states (Cape Verde, Comoros, Mauritius, Sao Tomé & Principé, and the Seychelles), assembled and published by the KEMRI Wellcome Trust Programme.^17^ The primary source of data were master facility lists (MFLs) of national Ministries of Health and documentation by United Nations and non-governmental organizations. Additional sources included websites and data portals by SSA governments, health sector reports, and personal communications. We henceforth refer to this dataset as the MFL dataset. 52% of the healthcare facilities contained in the data were manually geocoded by the KEMRI Wellcome Trust Programme team. For Sudan, Guinea-Bissau, and ten out of 18 provinces in Angola, the MFL dataset contains the geographic coordinates of hospitals only. The MFL dataset included 92,245 healthcare facilities in our study countries of which 4,720 were classified as hospitals. While the KEMRI Wellcome Trust Programme team used, among other tools, OSM to assign geocodes to healthcare facilities in the MFL dataset that had a missing geocode,^17^ they did not use OSM to identify healthcare facilities that were not already contained in the MFL dataset.

We verified the degree to which the GPS coordinates for a random sample of 20 healthcare facilities (320 facilities in total) for each of the 16 strata resulting from the possible permutations of healthcare facility type (primary care or hospital), dataset (OSM or MFL data), and region overlapped with building structures and human settlements in Bing satellite imagery. The results are shown in **Table S1**.

### Data source for the geolocation of the population

Population counts for adults aged 60 years and older were obtained from the WorldPop project.^22^ The counts reflect projections for 2020 at a spatial resolution of 1 km^2^. The WorldPop project built this dataset using a semi-automated dasymetric mapping method that employs a Random Forest classifier to disaggregate census data at the level of national census tracks to 1 km^2^ areas.^23^ Predictors used were geographical properties, such as topography, climate, and land cover, as well as the density of human-built features, such as nighttime lights, roads, and buildings.

### Estimating travel time to the nearest healthcare facility

We merged the OSM and MFL dataset such that our estimated travel times are the travel time to the nearest healthcare facility, regardless of the data source in which the facility is listed. We chose this strategy because both datasets are more likely to be missing existing healthcare facilities than to falsely list a non-existing healthcare facility. We estimated travel time to the nearest healthcare facility separately for hospitals and healthcare facilities of any type. Hospitals were chosen as one entity of interest because most healthcare interventions to care for individuals with severe COVID-19 require hospital-based care. Healthcare facilities of any type were chosen as an additional entity of interest because physical access to any healthcare facility likely influences the degree to which individuals with COVID-19 present to the healthcare system and, thus, the extent to which the healthcare system is made aware of new COVID-19 cases. In the absence of community-based screening for SARS-CoV-2 infections and ignoring that more remote areas may experience less SARS-CoV-2 transmission, areas with low physical access to healthcare facilities of any type may, thus, have a disproportionately high number of unreported COVID-19 cases.

We used the AccessMod tool (version 5.6.33) to estimate travel time.^25^ AccessMod employs a raster-based cost-distance algorithm, whereby each raster cell is associated with a cost value that determines the time required to travel through this cell. The cost for each cell was modelled using the 2018 Copernicus Global Land Cover product and the Shuttle Radar Topography Mission (SRTM v. 4) digital elevation model as basic impedance surface.^24,25^ In addition, we used OSM data to ascertain the road network and locations of rivers and open water (which were considered barriers for any kind of travel). Aligning with previous studies in SSA,^27^ we assigned a travel speed of 100km/h to motorways and primary roads, 50km/h to secondary roads, and 30 km/h to tertiary roads. Barren land and built-up areas were assigned a 5km/h and forests a 2 km/h walking speed. The model was created at a spatial resolution of 100m^2^. For both OSM and MFL data, we calculated the travel time from each cell to the nearest healthcare facility of any type and the closest hospital. These results were then aggregated to a 1km^2^ resolution to match the resolution of the WorldPop population data. Our analyses assumed that individuals were able to cross national borders to reach the nearest healthcare facility and we did not assign an additional time cost for a border crossing. Similarly, even though we show maps separately for each region, our travel time estimates assumed that individuals were able to cross regional borders and we did not apply a time cost for this crossing. We did not allow for variations in travel time by time of day or day of the week.

### Statistical analysis

We plotted the distribution of travel time, separately for hospitals and healthcare facilities of any type, in each country. In addition, we mapped the estimated travel time at a 1km x 1km resolution both as a continuous variable and when categorizing travel time into less than two hours, two to six hours, six to 12 hours, and more than 12 hours for the nearest hospital, and less than one hour, one to two hours, two to six hours, and more than six hours for the nearest healthcare facility of any type. When summarizing our data as binomial proportions, we show two-sided 95% confidence intervals using the Wilson score interval.^26^ Other than the calculation of travel time, which was carried out using AccessMod 5, all analyses were conducted in R version 3.6.3.^27^

## Results

### Sample characteristics

Across our two datasets, the population density of healthcare facilities varied from 0.067 facilities per 100,000 in Burkina Faso (MFL data) to 11.008 facilities per 100,000 in the Central African Republic (OSM data) for hospitals, and from 0.034 facilities per 100,000 in Eritrea (OSM data) to 28.053 facilities per 100,000 in Gabon (MFL data) for healthcare facilities of any type.

**Table 1.**
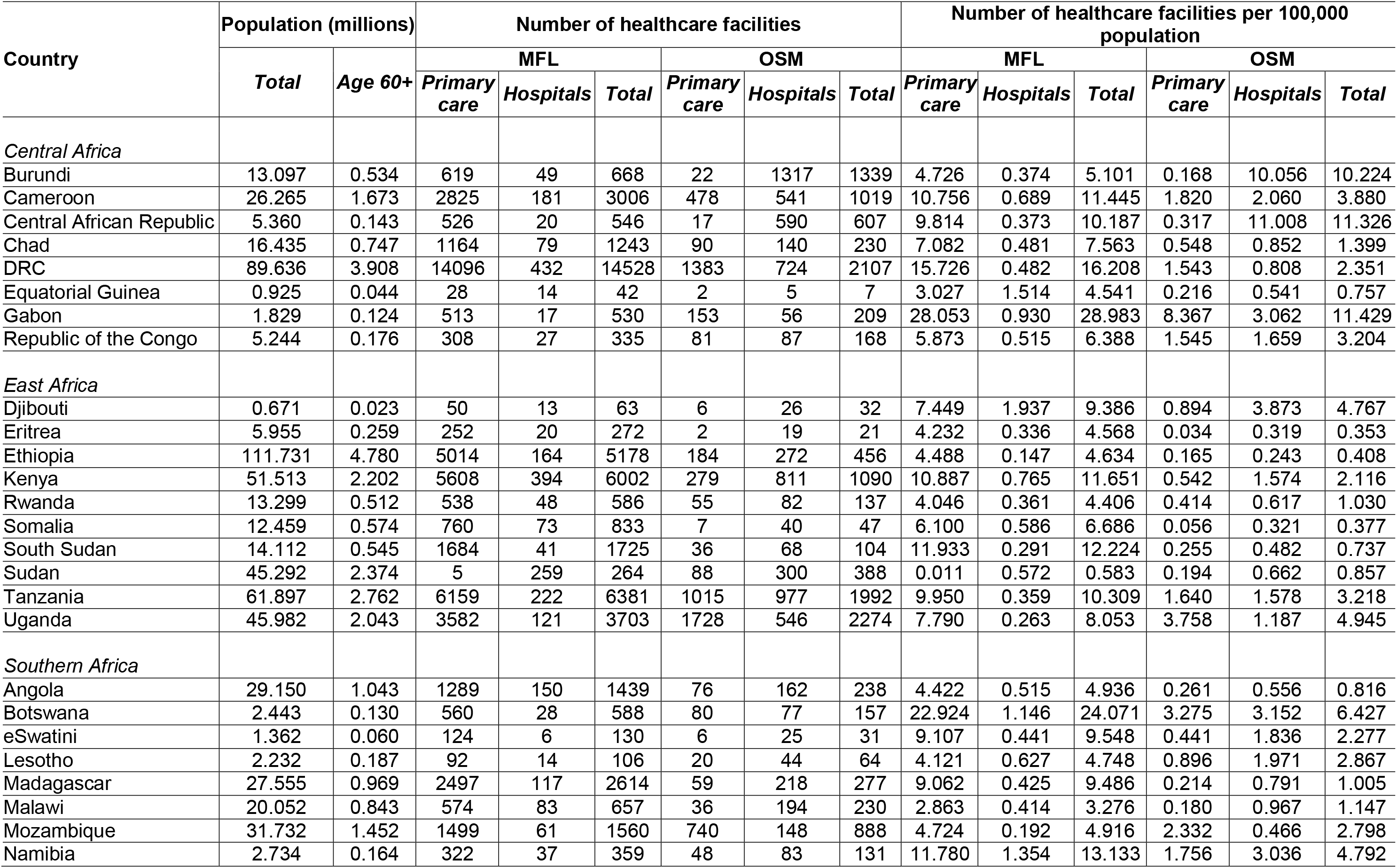
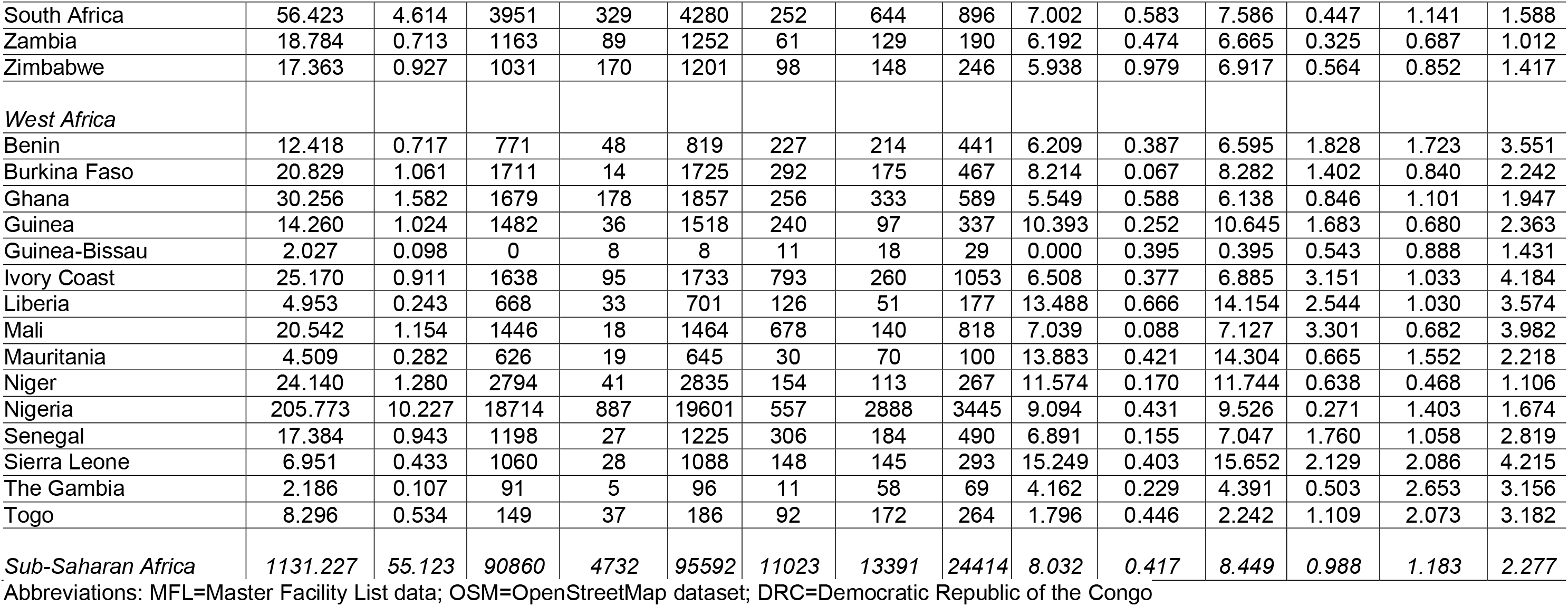
Population and number of healthcare facilities by country.

### Distribution of travel time to the nearest healthcare facility

Across SSA, the proportion of adults aged 60 years and older with an estimated travel time of greater than six hours to the nearest hospital was 9.6% (95% CI: 5.2 – 16.9) across SSA, varying from 0.0% (95% CI: 0.0 – 3.7) in Burundi and The Gambia to 40.9% (95% CI: 31.8 – 50.7) in Sudan (**Figure S1**). For healthcare facilities of any type and using a travel time cut-off of two hours, the corresponding proportions were 15.9% (95% CI: 10.1 – 24.4) across SSA, ranging from 0.4% (95% CI: 0.0 – 4.4) in Burundi to 59.4% (95% CI: 50.1 – 69.0) in Sudan (**Figure S2**).

The shape of the distribution of travel time to the nearest hospital for adults aged 60 years and older varied greatly across countries (**Figure 1**). It ranged from a distribution in which the vast majority of the population is within 60 minutes travel time (e.g., in Burundi), to distributions in which the population was almost equally spread across the range of travel time from 0 minutes to four hours (e.g., in Ethiopia). For the nearest healthcare facility of any type, in contrast, the distribution was more heavily skewed towards very short travel times (**Figure 2**), with the proportion of adults aged 60 years and older who reside within 30 minutes of traveling to the nearest facility being at least 25% in 43 of our 44 study countries. The travel time distributions are shown separately for the MFL and the OSM dataset in **Figures S3 to S6**.

**Figure 1.**
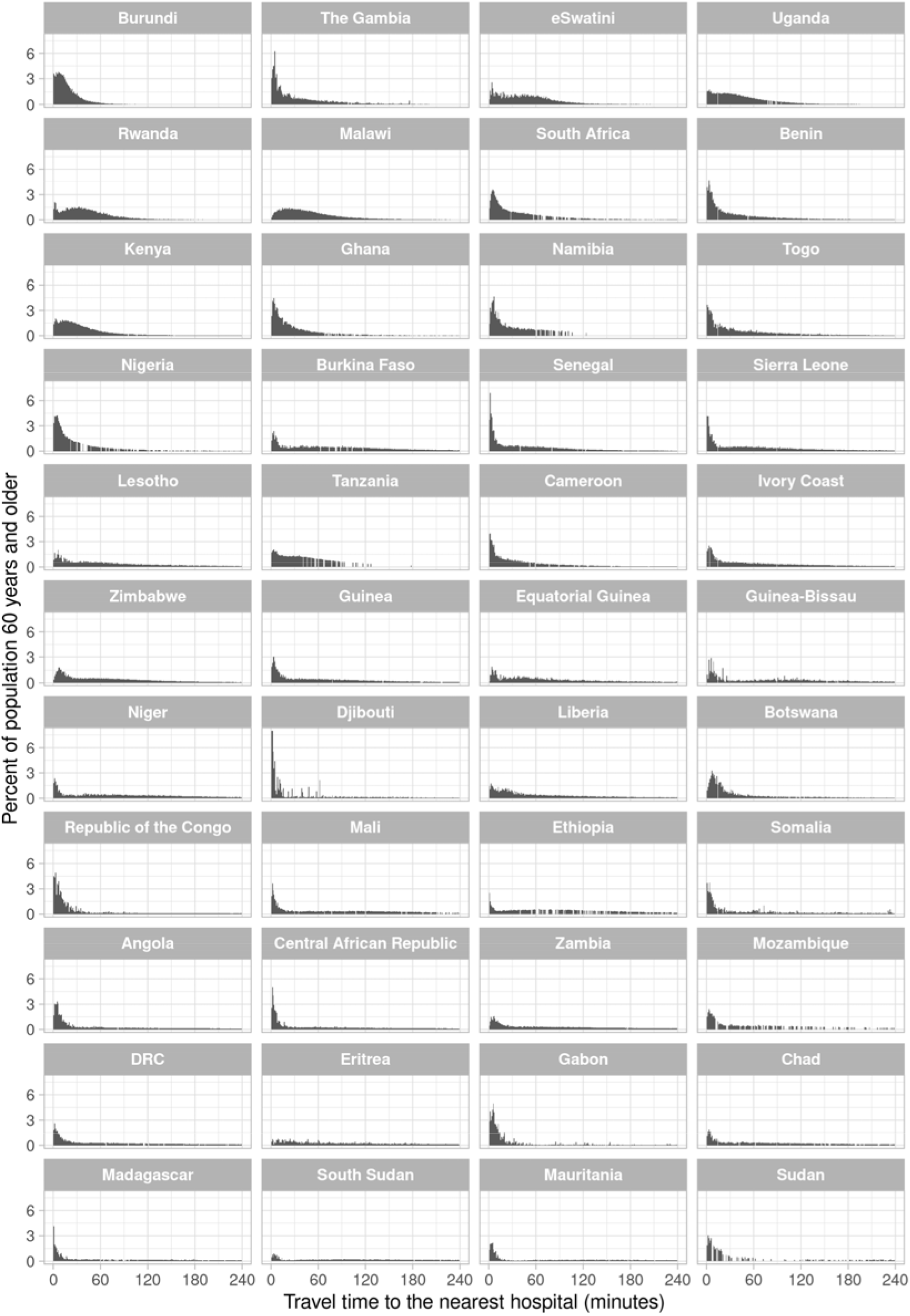
Distribution of travel time to the nearest hospital for adults aged 60 years and older, by country^1^. Abbreviations: DRC=Democratic Republic of the Congo ^1^ Countries were ordered in ascending order by the proportion of adults aged 60 years and older in their population who reside in a 1km x 1km area that has an estimated travel time >6 hours to the nearest hospital.

**Figure 2.**
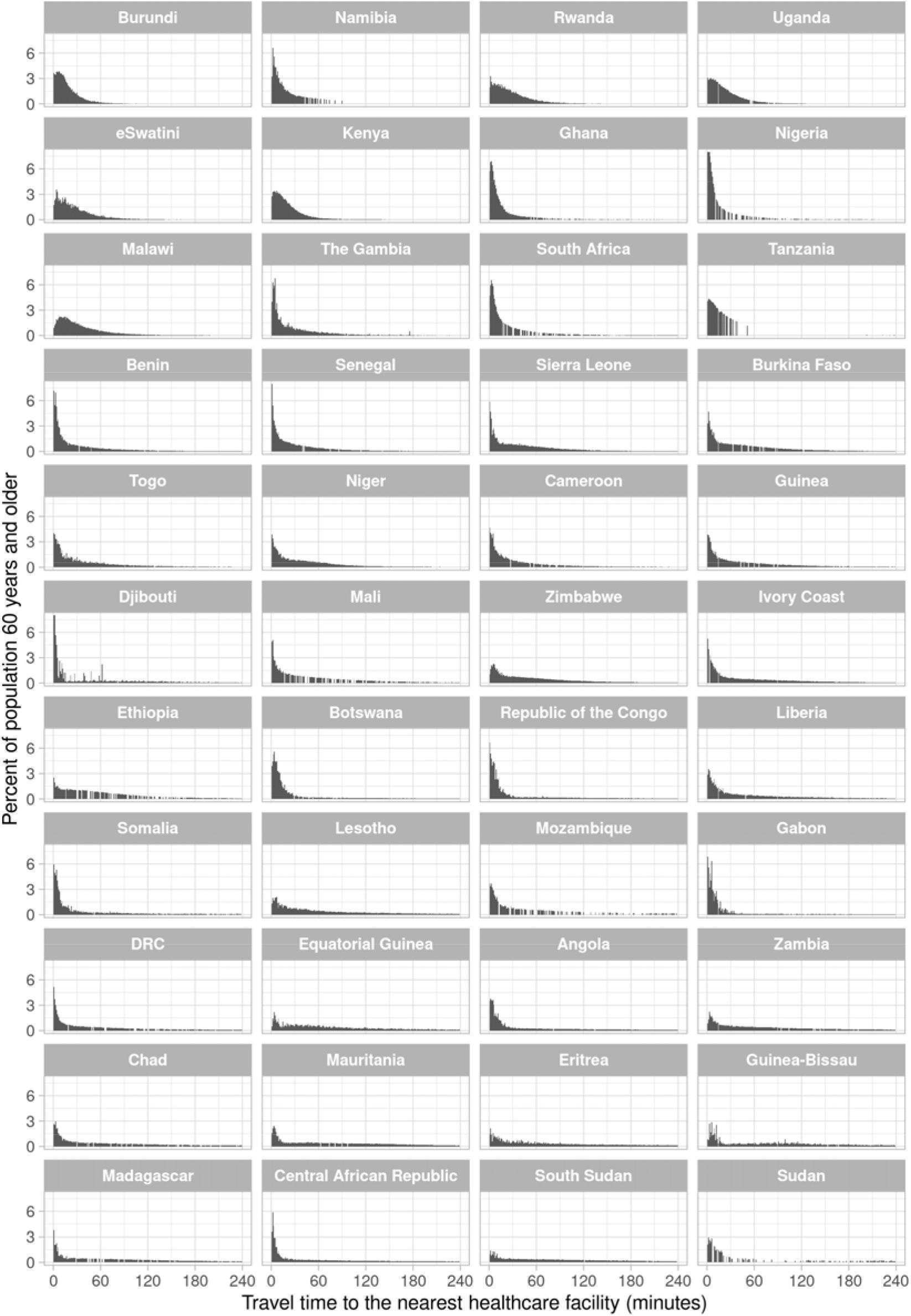
Distribution of travel time to the nearest healthcare facility of any type for adults aged 60 years and older, by country^1^. Abbreviations: DRC=Democratic Republic of the Congo ^1^ Countries were ordered in ascending order by the proportion of adults aged 60 years and older in their population who reside in a 1km x 1km area that has an estimated travel time >2 hours to the nearest healthcare facility.

### Maps of travel time to the nearest hospital

**Figure 3** shows the population density of adults aged 60 years and older as well as the estimated travel time among these adults to the nearest hospital at a 1km x 1km resolution. The third column of maps focusses on populated areas (which we defined as areas with at least one adult aged 60 years and older per km^2^) and categorizes travel time into less than two hours, two hours to less than six hours, six hours to less than 12 hours, and more than 12 hours. This column shows that almost all countries in SSA contain populated areas that have an estimated travel time to the nearest hospital of greater than 12 hours (indicated as the areas in dark red). Countries with many of these 1km^2^ areas with poor physical access to hospital care included the Democratic Republic of the Congo, Madagascar, Ethiopia, Sudan, South Sudan, Mozambique, and Mauritania. More detailed maps created separately for each country are shown in **Figures S7 – S50**. Regional maps created using only the MFL and only the OSM data are shown in **Figure S51** and **Figure S52**, respectively.

**Figure 3.**
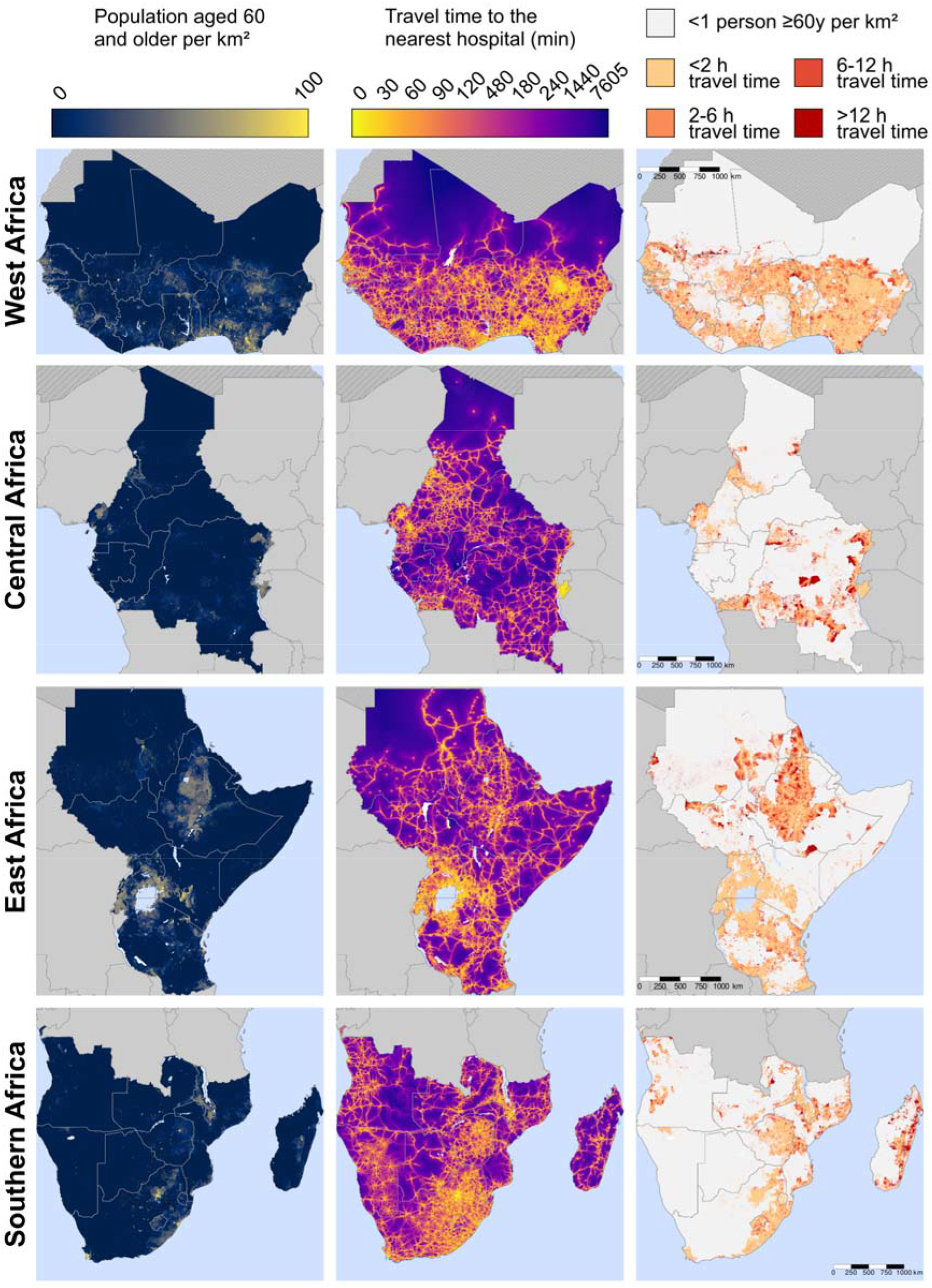
Maps of travel time to the nearest hospital for adults ≥60 years, by region.

### Maps of travel time to the nearest healthcare facility of any type

**Figure 4** shows the same variables as Figure 3 but for healthcare facilities of any type (as opposed to hospitals only). Countries with a high number of these 1km^2^ areas with poor physical access to a healthcare facility included the Democratic Republic of the Congo, Sudan, Ethiopia, Madagascar, Mozambique, South Sudan, and Angola. Maps created separately for each country are shown in **Figures S53 – S96**. Regional maps created using only the MFL and only the OSM data are shown in **Figure S97** and **Figure S98**, respectively.

**Figure 4.**
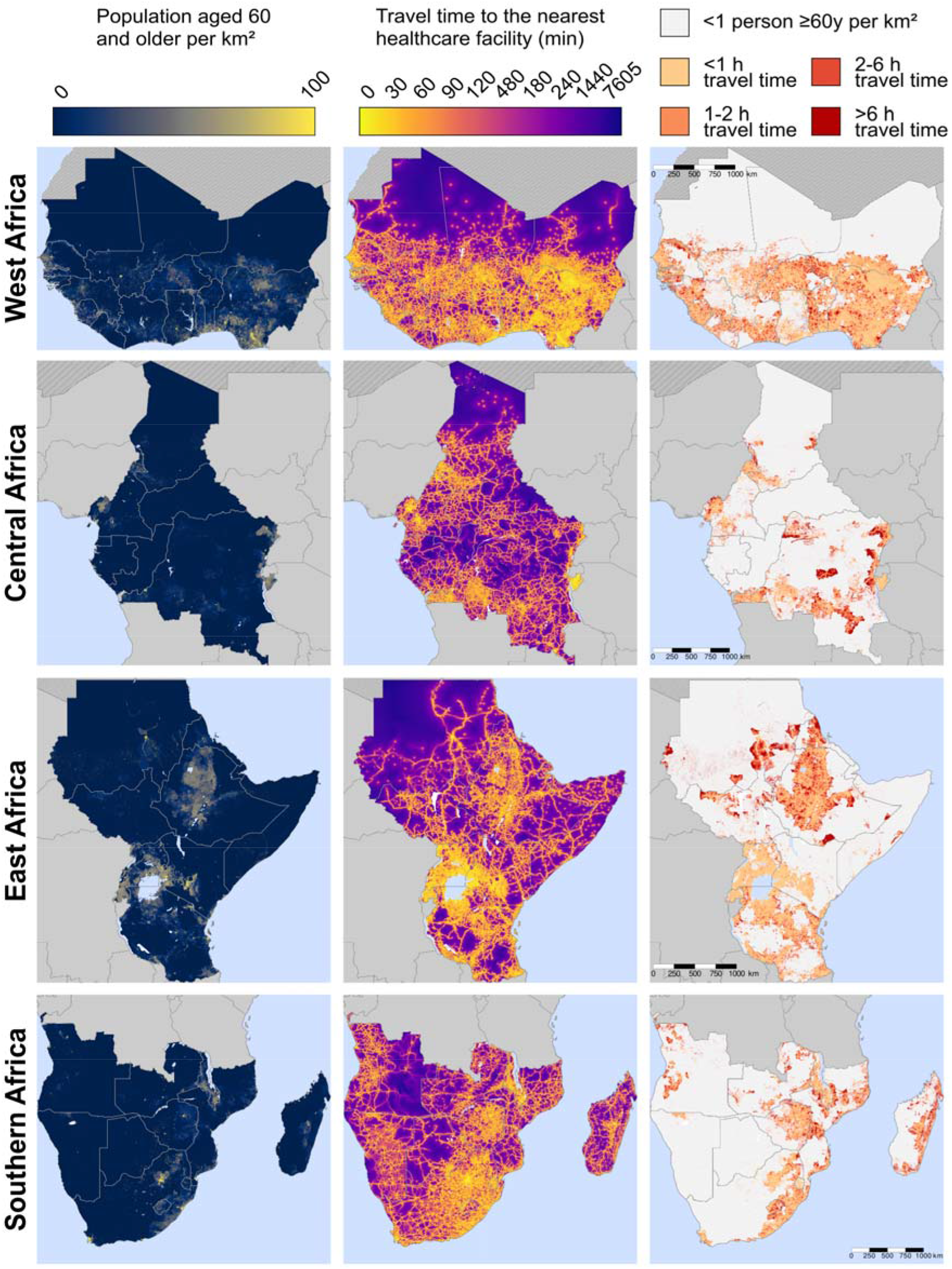
Maps of travel time to the nearest healthcare facility for adults ≥60 years, by region.

## Discussion

With approximately 10% of adults aged 60 years and older across SSA having an estimated travel time to the nearest hospital of greater than six hours, physical access to healthcare will likely play a major role in whether older adults in this world region will be able to seek care for COVID-19. By precisely identifying where older adults are residing who have a particularly high estimated travel time to the nearest hospital, our high-resolution maps can inform policy makers as to where interventions to increase physical access to hospital care are needed most urgently. Such interventions could include transport programs to existing hospitals and the establishment of make-shift hospitals. In addition, our maps of estimated travel time to the nearest healthcare facility of any type could help guide policy makers as to which populations are least likely to present to the healthcare system when they suffer from COVID-19 symptoms due to a lack of physical access to healthcare. This information in turn could be helpful in interpreting monitoring data on new cases of COVID-19 from different areas within countries, and in targeting testing efforts to those populations that have the greatest need for such tests.

The usefulness and policy relevance of this analysis goes beyond informing countries’ response to the SARS-CoV-2 pandemic. Physical access – that is, the time required to travel to a healthcare facility, available transport options, and costs for transport – is thought to be one of the main barriers to accessing healthcare in SSA.^5-10^ Yet, there is currently very little detailed evidence on how physical access to healthcare varies across SSA, particularly within countries. Such evidence, however, is crucial to guide policy makers in identifying those areas that have the greatest need for community outreach programs, the establishment of new healthcare facilities, and improved transport infrastructure. Our study helps fill this important evidence gap for older adults in the region and is, thus, of high relevance for informing countries’ efforts to improve care for conditions that affect older adults, particularly chronic non-communicable diseases. Specifically, this study builds on the, to our knowledge, four existing studies that have mapped physical access to healthcare in SSA at a subnational level within countries, as well as studies that have examined physical access to man-made resources more generally.^28^ Ouma et al. have examined access to emergency hospital care in SSA.^29^ This study differs from ours in that it focussed on women of child-bearing age (15 to 49 years) rather than older adults, did not include primary healthcare facilities nor any private-sector healthcare facilities, did not use OSM data, employed a cut-off of travel time less or greater than two hours (based on a target set by the Lancet Commissions for Global Surgery 2030^30^) rather than analysing the whole distribution, analysed data from 2015, and did not provide detailed country-by-country maps. Other relevant studies have focussed on the effect of physical access to a healthcare facility on the probability of seeking care for a febrile episode in children,^31^ estimated travel time to healthcare facilities among populations at risk of viral haemorrhagic fevers,^32^ and physical access to major district and regional hospitals.^33^

Another key contribution of our study is the collation of a new dataset on geo-tagged healthcare facilities in SSA. By making this dataset available in the public domain and including the location of other age groups (not merely adults aged 60 years and older), we allow researchers and policy makers to run their own analyses for a variety of demographic groups and add to (or alter) the list of geo-tagged healthcare facilities in a country. There is currently no authoritative source of the location of all healthcare facilities in SSA. We have combined data from the only two existing sources of data for the geolocation of healthcare facilities in the region. We chose this approach because it is highly likely that neither dataset is complete, as is evidenced by the fact that in some countries the MFL dataset listed a higher number of healthcare facilities than the OSM data while the opposite was the case in other countries. Because the OSM project relies on volunteers to map and tag healthcare facilities, the OSM data by itself is particularly likely to underestimate the density of healthcare facilities in an area. In addition, because the categorization of a healthcare facility as a primary care facility or a hospital relies on the judgement or knowledge of the person tagging the facility, the OSM data likely has inaccuracies in the categorization of healthcare facilities into different types. For instance, OSM listed far more hospitals than primary care facilities for the Central Africa Republic, which appears unlikely to be correct. The fact that OSM contained a higher number of healthcare facilities in many countries than did the MFL dataset, especially for hospitals, is, however, encouraging in that OSM appears to be a useful source of information for the geolocation of healthcare facilities. Importantly, OSM data are likely to improve over time as coverage of smartphones increases in SSA and more volunteers map out their local areas. We will, therefore, update our dataset on a regular basis. Moving forward, it will be important to continuously monitor the validity of the data entered into the OSM and MFL dataset; a task that would ideally be accomplished by Ministries of Health in SSA.

This study has several limitations. First and foremost, while we (by combining MFL with OSM data) likely provide the most comprehensive source of data for the geolocation of healthcare facilities that is currently available, it is still likely to miss a substantial proportion of healthcare facilities. The degree to which this is the case probably varies between countries as both the participation in the OSM project and the degree to which documentation used for the MFL dataset were available and complete differ across countries.^34^ Second, we do not have any data on the readiness of healthcare facilities to provide care, nor the quality of care provided at healthcare facilities. Similarly, we did not have information on the functioning of referral systems from primary to secondary and tertiary care, which impacts access to effective healthcare for COVID-19 and other conditions requiring more specialized care. These factors are likely to vary across and within countries.^35^ Third, a limitation of our analysis for the COVID-19 response is that governments may decide that not all hospitals in a country should be providing care for COVID-19. Fourth, our analysis does not take into account that vulnerability to COVID-19 is likely affected by factors beyond age that vary across and within countries, including HIV, tuberculosis, and malnutrition. We decided against including these factors in our analysis because which conditions increase the risk of experiencing a severe disease course, and to what degree, is still largely unknown, especially in SSA. Fifth, we did not examine to what degree the MFL and OSM dataset contain the same healthcare facilities. Our findings are, thus, estimates for travel time to the nearest healthcare facility regardless of whether the facility is contained in the MFL or the OSM dataset. This strategy does not introduce any bias as long as the same healthcare facility has the same or very similar geographic coordinates in both datasets. It is, however, possible that the geographic coordinates for the same healthcare facility differed between the two datasets, in which case our analysis would consider these to be two different healthcare facilities and, thus, underestimate the true travel time. Lastly, our travel time numbers are approximations that, for example, do not take into account the frequency of transport services and assign an estimated (rather than measured) travel speed to different types of roads. Similarly, we assumed that individuals were able to cross national borders and incurred no additional time cost from doing so. In border regions where these assumptions do not hold true, our estimated travel times would, thus, underestimate the true travel time. Lastly, our analysis focusses on only one aspect of access to healthcare and thus does not, for instance, take into account financial barriers to accessing care.

Most countries in SSA contain areas in which older adults have little to no physical access to a hospital and (albeit to a lesser extent) healthcare facilities of any type. If COVID-19 becomes a generalized epidemic that infects large swaths of countries’ populations in the region, then it will be older adults living in these areas who are in a particularly high need for either improved transport options to existing hospitals or the provision of make-shift hospital care. Beyond their usefulness for the COVID-19 response, our maps could inform health system planning for conditions that commonly affect older adults, such as expansions of care for myocardial infarctions and strokes.

## Data Availability

The data that support the findings of this study are openly available in heiData at https://doi.org/10.11588/data/RGM2AW.

## Contributors

PG wrote the first draft of the manuscript. MR conducted the data analysis. PG, MR, SL, and AZ conceptualized the study. All authors provided crucial input on several iterations of the manuscript and approved the final version.

## Acknowledgements

The authors wish to acknowledge the contribution of scientists at the KEMRI-Wellcome Trust Programme who assembled the MFL dataset since 2010 including Robert Snow, Peter Macharia, and Joseph Maina.

